# Prevalence and Determinants of Academic Bullying Among Junior Doctors in Sierra Leone

**DOI:** 10.1101/2024.11.13.24317261

**Authors:** Fatima Jalloh, Ahmed Tejan Bah, Alieu Kanu, Mohamed Jan Jalloh, Kehinde Agboola, Monalisa M.J. Faulkner, Foray M. Foray, Onome T. Abiri, Arthur Sillah, Aiah Lebbie, Mohamed B. Jalloh

## Abstract

**Background:** Academic bullying among junior doctors—characterized by repeated actions that undermine confidence, reputation, and career progression—is associated with adverse consequences for mental health and professional development. We aimed to investigate the prevalence and determinants of academic bullying among junior doctors in Sierra Leone.

**Methods:** We conducted a cross-sectional survey of 126 junior doctors at the University of Sierra Leone Teaching Hospitals Complex (USLTHC) in Freetown between January 1 and March 30, 2024. Participants were selected through random sampling. Data were collected using a semi-structured, self-administered questionnaire and analyzed with descriptive statistics and multivariable logistic regression.

**Results:** Of the 126 participants (61.1% male; mean age 31.9 years), 86 (68.3%) reported experiencing academic bullying. Among those, 54.6% experienced it occasionally and 35.2% very frequently. The most common forms were unfair criticism (73.3%), verbal aggression (66.3%), and derogatory remarks (47.7%). Consultants and senior doctors were the main perpetrators, with incidents primarily occurring during ward rounds, clinical meetings, and academic seminars. No statistically significant predictors of bullying were found for gender (odds ratio [OR] 2.07, 95% CI 0.92–4.64; p=0.08) or less than two years of practice (OR 0.30, 95% CI 0.05–1.79; p=0.19).

**Conclusion:** Academic bullying is pervasive among junior doctors at USLTHC, with significant implications for their mental health and professional development. Urgent implementation of comprehensive strategies—including culturally sensitive policies, targeted training programs, confidential reporting mechanisms, and leadership development—is essential to address this issue. Promoting ethical leadership and fostering a culture of respect may mitigate incivility and burnout, enhancing the work environment for junior doctors.

## Introduction

Academic bullying—defined as maltreatment within academic settings intended to hinder the professional or academic progress of targeted individuals—remains a pervasive issue in medicine, particularly affecting junior doctors.(1)

The hierarchical and demanding nature of the medical profession has historically facilitated such behavior, leading to significant implications for mental health, professional interactions, and career advancement.(1,2) Common manifestations include public humiliation, verbal abuse, micromanagement, excessive workload, exclusion, sabotage, gaslighting, and undermining.(3) Despite these adverse effects, cultural norms that emphasize respect for authority figures often create barriers to reporting and intervention; victims may be reluctant to speak out against superiors.(4)

Studies in other contexts have shown that a significant number of junior doctors experience bullying during their training, including overt verbal abuse, humiliation, excessive criticism, and exclusion from learning opportunities.(1,2,5,6) The persistent perpetration of these behaviors fosters a detrimental work environment that adversely affects the well-being and career prospects of junior doctors.(1)

Factors such as favoritism, nepotism, and limited resources can exacerbate academic bullying by fostering resentment and competition while restricting access to learning opportunities.(1,2) Victims are more prone to anxiety, depression, exhaustion, decreased job satisfaction, and may consider leaving the profession.(6,7) Furthermore, academic bullying can negatively impact patient safety and contribute to long-term academic and psychiatric issues.(8) This culture of mistreatment not only affects individual well-being but also undermines team functioning and healthcare delivery.(9,10)

Given Sierra Leone’s healthcare infrastructure challenges and workforce shortages,(11) the additional strain of academic bullying on junior doctors is particularly concerning. While studies elsewhere report bullying rates as high as 83%,(2,5,12) there is a lack of data on its prevalence and consequences in Sierra Leone. Therefore, we aimed to investigate the prevalence and determinants of academic bullying among junior doctors at the University of Sierra Leone Teaching Hospitals Complex (USLTHC) in Freetown, Sierra Leone.

## Methods

### Study design and setting

We conducted a cross-sectional survey at the major hospitals of USLTHC in Freetown, Sierra Leone. The USLTHC - Connaught Hospital, Princess Christian Maternity Hospital, Ola During Children’s Hospital, and Sierra Leone Psychiatry Teaching Hospital are the largest and primary government referral hospitals in the country and serve as the main training centers for junior doctors, including registrars (residents) and house officers (interns) in Sierra Leone. The survey was conducted from 1st January to 30th March 2024.

### Participants and sampling

#### Inclusion and exclusion criteria

All junior doctors who had been employed for a period of six months or longer and had reached the age of 18 years or older were included in the study. Those who were on outside posting or leave (annual or sick) were excluded, and no visiting junior doctors outside of USLTHC were included. The 6-month working experience requirement was used as the cutoff to ensure that participants have had sufficient interaction with both superiors and contemporaries during their training or postings.

#### Sampling strategy and sample size

A random sampling strategy was employed, drawing from a list of junior doctors over the age of 18 who had been employed at the University of Sierra Leone Teaching Hospitals Complex (USLTHC) for at least six months. This list constituted our sampling frame. To determine the appropriate sample size for the study, we utilized the Yamane formula for sample size estimation in cross-sectional research: **n = *N* / [1 + N(e^2^)])**, where **n** is the required sample size, ***N*** is the total population size, and **e** is the margin of error set at 5% (0.05).(13) Given an estimated population of 160 eligible junior doctors, the formula yielded a sample size of 114. Anticipating potential non-response or incomplete data, we added an additional 10% to this figure, resulting in a final sample size of 126 participants. These participants were drawn from various departments across the four sites of the USLTHC.

#### Data collection instrument

Data were collected using a semi-structured, self-administered questionnaire. To accommodate participants who might not have had time to complete the paper-based version, the survey was also made available online via a secure server using Microsoft Forms. The questionnaire captured demographic information, including sex, age, duration of practice or training, and job title or designation.

The primary outcome measure was the respondent’s experience of workplace bullying, determined by a yes or no response to the question: “Have you experienced any form of workplace bullying in the last six months while training?”

Respondents who indicated they had experienced bullying were asked to describe the nature of the bullying encountered. Bullying was defined as any instance of intimidation, humiliation, degradation, misuse of power, or abuse of position or authority that caused the respondent to feel defenseless and undermined their sense of dignity.(1,2,14) Information was collected on whether respondents had experienced any such acts in the past six months. The survey instrument was developed and piloted with ten participants to ensure clarity and relevance, with refinements made based on their feedback.

#### Statistical analysis

We conducted descriptive statistics to summarize the data. For continuous variables with a normal distribution, we reported means and standard deviations; for non-normally distributed variables, medians and interquartile ranges were provided. Associations between categorical variables were assessed using Pearson’s chi-squared tests or Fisher’s exact tests, as appropriate. Results were presented in tables and graphical summaries.

To explore independent associations between pre-specified characteristics and the primary outcome—respondents’ experience of workplace bullying—we performed multivariable logistic regression analyses. Explanatory variables were selected based on their relevance and included age (≤34 years vs. ≥35 years), sex (male vs. female), marital status (married vs. others), level of training (house officer and others vs. registrar), and duration of practice (≤2 years vs. ≥3 years). Results were reported as odds ratios (ORs) with 95% confidence intervals (CIs) and corresponding p-values. Statistical significance was set at a 5% level. All analyses were conducted using the Statistical Package for the Social Sciences (SPSS) version 27.

#### Participant and public involvement statement

Due to unexpected delays and time constraints, we were unable to involve participants or the public in the study’s design, execution, or reporting. However, we are now considering a higher level of public and stakeholder engagement when sharing our research findings.

#### Ethical considerations

The study received ethical approval from the College of Medicine and Allied Health Sciences Institutional Review Board (IRB review number: COMAHS/IRB/013-2024). Informed consent was obtained from all participants at the outset of the questionnaire. All responses were anonymous and confidential, adhering to established ethical standards. No personally identifiable information was collected, and all necessary measures were taken to ensure proper storage and management of the data to maintain confidentiality and data integrity.

## Results

### Socio-demographic characteristics of participants

A total of 126 individuals completed the survey, comprising 77 males (61.1%) and 49 females (38.9%). The mean age of the participants was 31.9 years, with a standard deviation (SD) of 5.05 years. Regarding marital status, 68 individuals (53.9%) were single and never married, 52 (41.3%) were married or in a domestic partnership, 2 (1.6%) were separated, 1 (0.8%) divorced, and 3 (2.4%) preferred not to disclose their marital status (**Table 1**).

**Table 1.** Socio-demographic characteristics of the respondents n =126.

| Characteristics | Frequency (%) |
| --- | --- |
| Age (years) |  |
| 18-24 | 2 (1.6) |
| 25-34 | 95 (75.4) |
| 35-44 | 26 (20.6) |
| 45-54 | 3 (2.4) |
| Mean age (SD) | 31.9 (5.05) |
| Sex |  |
| Female | 49 (38.9) |
| Male | 77 (61.1) |
| Marital Status |  |
| Single, never married | 68 (53.9) |
| Married or domestic partnership | 52 (41.3) |
| Separated | 2 (1.6) |
| Divorced | 1 (0.8) |
| Prefer not to say | 3 (2.4) |
| Level of training |  |
| House Officer | 59 (46.8) |
| Medical Officer | 22 (17.5) |
| Registrar | 43 (34.1) |
| Senior Registrar | 2 (1.6) |
| Duration of practice (years) |  |
| < 2 | 66 (52.4) |
| 2 and above | 60 (47.6) |

| Current Training Department |  |
| --- | --- |
| Internal Medicine | 35 (27.8) |
| Surgery and its sub-specialties | 34 (26.9) |
| Paediatrics | 21 (16.7) |
| Obstetrics & Gynaecology | 23 (18.3) |
| Family Medicine | 5 (3.9) |
| Psychiatry | 6 (4.8) |
| Laboratory Medicine | 2 (1.6) |

In terms of level of training, the sample included 59 house officers (46.8%), 22 medical officers (17.5%), 43 registrars (34.1%), and 2 senior registrars (1.6%). The duration of practice varied, with 66 participants (52.4%) having practiced for 2 years or less and 60 participants (47.6%) having practiced for 3 years or more (**Table 1**).

Participants were also categorized by their current training departments. Internal Medicine had the highest representation, with 35 individuals (27.8%), followed by Surgery and its sub-specialties with 34 individuals (26.9%). Paediatrics included 21 participants (16.7%), Obstetrics & Gynaecology had 23 participants (18.3%), Family Medicine included 5 participants (3.9%), Psychiatry had 6 participants (4.8%), and Laboratory Medicine included 2 participants (1.6%) (**Table 1**).

This study examined the prevalence and forms of academic bullying among 126 participants. A total of 86 individuals (68.3%) reported experiencing bullying, while 40 individuals (31.8%) did not report such experiences (**Table 2**). Among the participants who reported being bullied, 48 (54.6 %) experienced bullying occasionally, and more than one-third (35.2%) experienced bullying very frequently (**Figure 1**).

**Figure 1.**
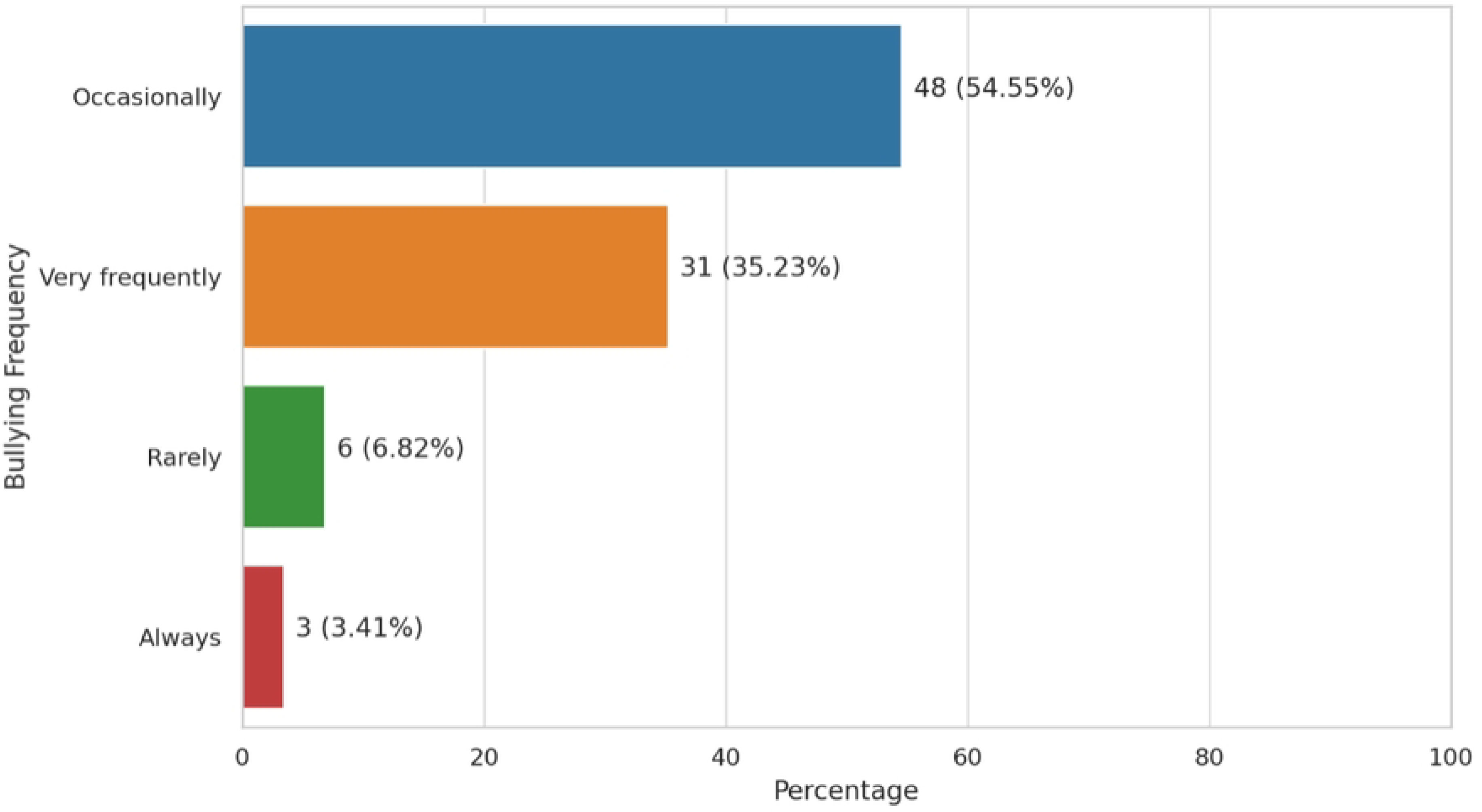
Frequency of bullying experienced by junior doctors.

**Table 2.** Prevalence and forms of academic bullying.

| **Variable** | **Frequency (%)** |
| --- | --- |
| **Current experience of bullying (n=126)** |  |
| Experienced | 86 (68.3) |
| Not Experienced | 40 (31.8) |
| **\*Forms of bullying (n=86)** |  |
| Unfair criticism or evaluation | 63 (73.3) |
| Verbal aggression | 57 (66.3) |
| Derogatory remarks | 41 (47.7) |
| Threat or intimidation | 33 (38.4) |
| Undermining dignity at work | 30 (34.9) |
| Exclusion from academic activities | 16 (18.6) |
| Others (extra on-call service) | 1 (1.2) |
| **#Common perpetrators of bullying (n=86)** |  |
| Consultants | 72 (83.7) |
| Other senior doctors (colleagues) | 66 (76.7) |
| Nursing staff | 18 (20.9) |
| Administrative staff | 15 (17.4) |
| Peers | 13 (15.1) |

\*Percentages are calculated based on the total number of respondents who reported any form of bullying or reported any type of perpetrator (n=86).

Among those who reported experiencing bullying (n=86), the most common forms of bullying included unfair criticism or evaluation, reported by 63 individuals (73.3%), and verbal aggression, reported by 57 individuals (66.3%). Derogatory remarks were reported by 41 individuals (47.7%), and threats or intimidation were experienced by 33 individuals (38.4%). Other reported forms of bullying included undermining dignity at work (30 individuals, 34.9%), exclusion from academic activities (16 individuals, 18.6%), and extra on-call service demands (1 individual, 1.2%) (**Table 2**).

Regarding the common perpetrators of bullying (n=86), consultants were identified as the most frequent perpetrators, reported by 72 individuals (83.7%). Other senior doctors were reported by 66 individuals (76.7%) as perpetrators. Additionally, 18 individuals (20.9%) reported nursing staff as perpetrators, 15 individuals (17.4%) reported administrative staff, and 13 individuals (15.1%) reported peers as perpetrators of bullying (**Table 2**).

The most common context or setting in which academic bullying occurred was during ward rounds, reported by 73 participants (82.95%). Clinical meetings were another context in which 51 individuals (57.95%) experienced bullying. Fifty individuals (56.82 %) reported academic seminars or presentations as the context for bullying. Lastly, administrative meetings were identified as a bullying setting by eight individuals (9.09%) (**Figure 2**).

**Figure 2.**
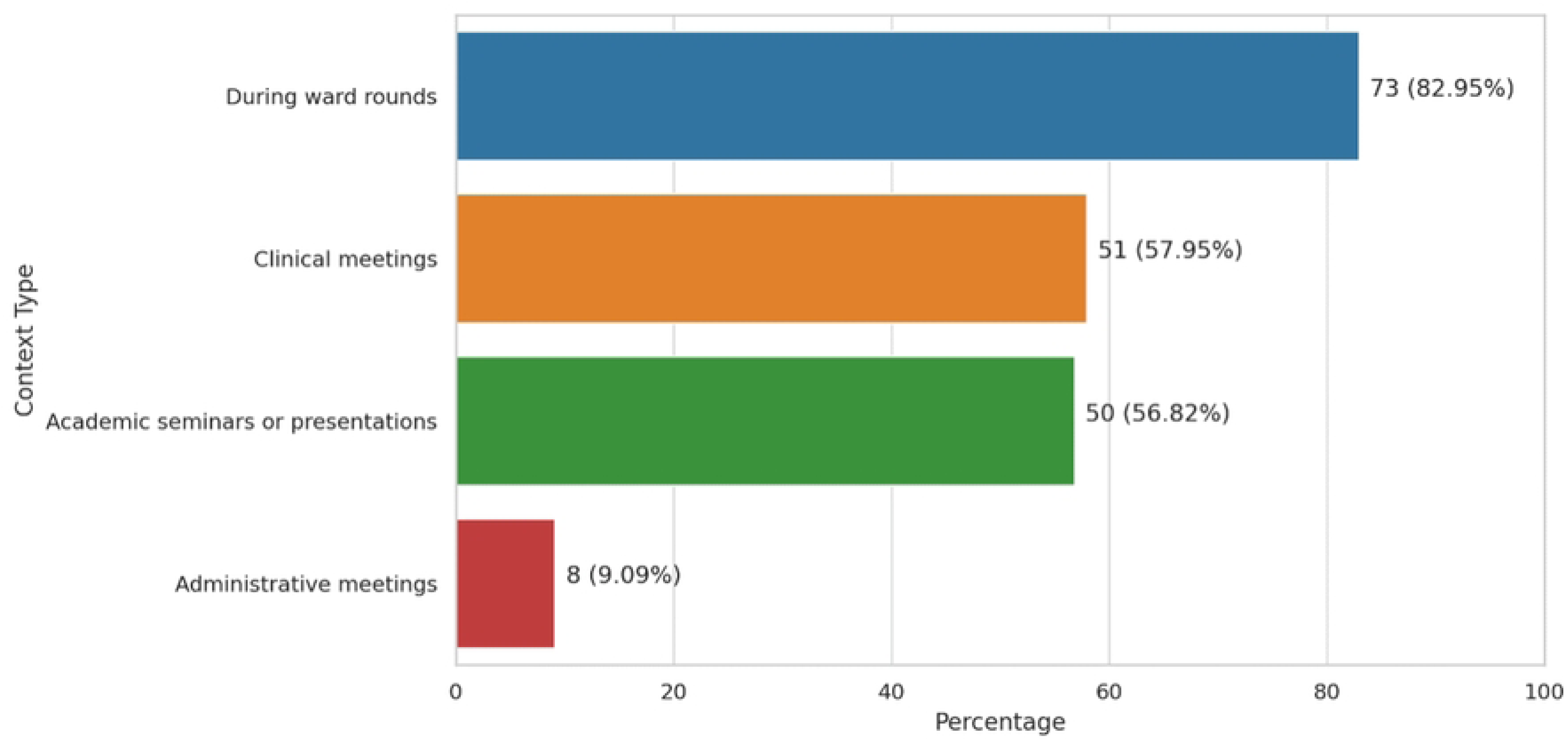
Context or setting of bullying activity.

### Multiple logistic regression analysis of factors independently associated with bullying

The logistic regression analysis did not identify any statistically significant predictors of bullying at the 5% significance level. Participants aged ≥35 years had 0.78 times the odds of experiencing bullying compared to those aged ≤34 years (OR = 0.78, 95% CI: 0.29 to 2.14, p = 0.63). House officers had 0.66 times the odds of experiencing bullying compared to registrars (OR = 0.66, 95% CI: 0.10 to 4.34, p = 0.67), while participants in the “Others” designation category (medical officers and senior registrars) had 2.58 times the odds of experiencing bullying compared to registrars (OR = 2.58, 95% CI: 0.67 to 9.92, p = 0.17). Marital status showed that participants categorized as “Others” had 0.94 times the odds of experiencing bullying compared to married or domestic partnership participants (OR = 0.94, 95% CI: 0.38 to 2.35, p = 0.90) (**Table 3**).

**Table 3.**
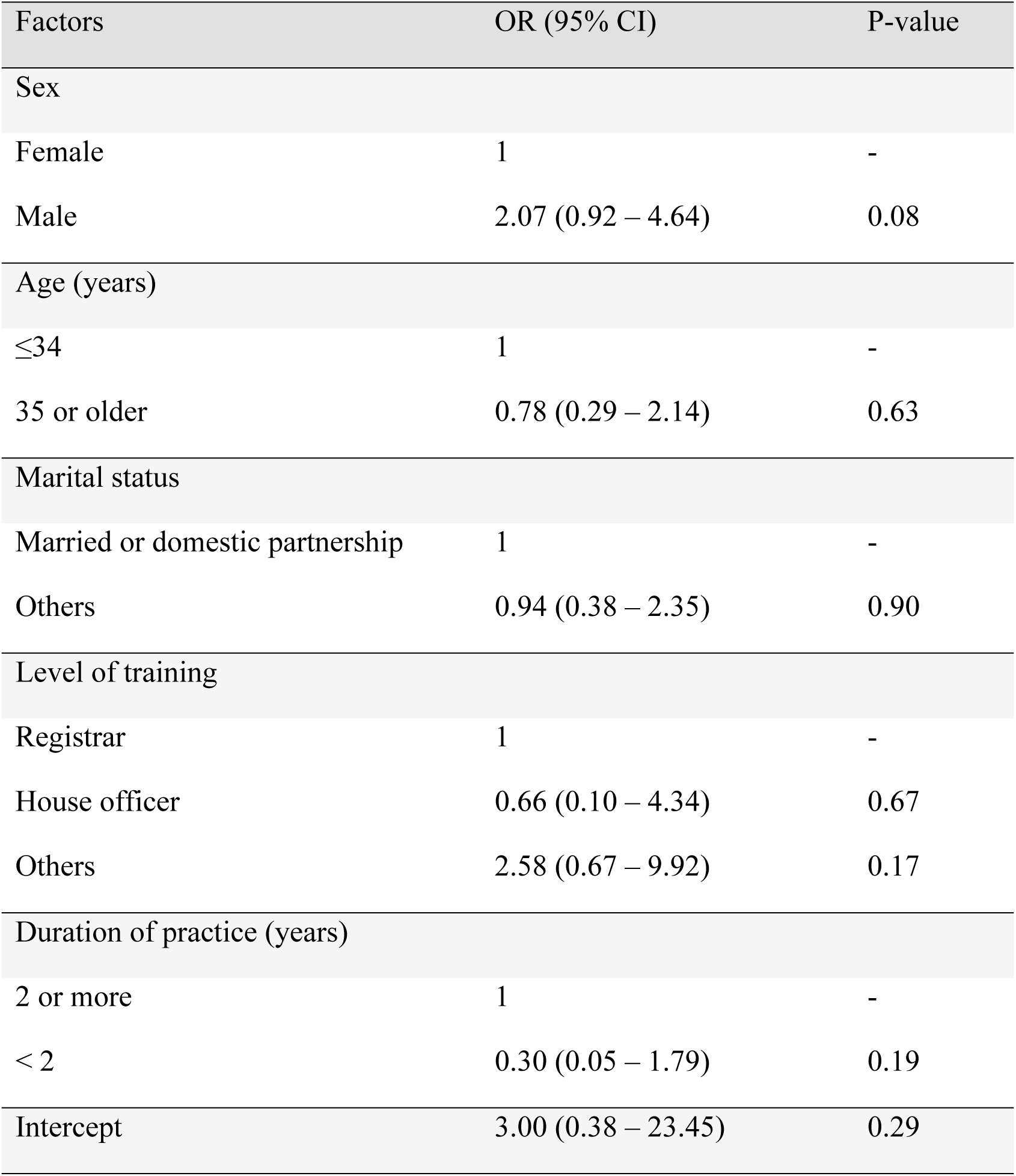
Multiple logistic regression analysis of factors independently associated with bullying.

Males had 2.07 times the odds of experiencing bullying compared to females (OR = 2.07, 95% CI: 0.92 to 4.64, p = 0.08). Participants with <2 years of practice had 0.30 times the odds of experiencing bullying compared to those with more than 2 years of practice (OR = 0.30, 95% CI: 0.05 to 1.79, p = 0.19) (**Table 3**).

The intercept, representing the log odds of experiencing bullying for the reference category (≤34 years old, female, married, registrar, ≥3 years of practice), had an odds ratio of 3.00 (95% CI: 0.38 to 23.45, p = 0.29), which serves as the baseline for comparison but is not directly interpretable in the same way as the other predictors (**Table 3**).

## Discussion

In this cross-sectional study, we investigated the prevalence and determinants of academic bullying among junior doctors at USLTHC in Freetown, Sierra Leone between 1st January and March 30, 2024. We found a high prevalence of bullying (68.3 %) among 126 participants, with unfair criticism and verbal aggression being the most common forms. Consultants and other senior doctors were frequently identified as perpetrators. Bullying occurred most frequently during ward rounds and clinical meetings. Despite the high prevalence, the analysis did not find any factors that were significantly associated with the likelihood of experiencing bullying.

The high prevalence of academic bullying in this study is much higher than the global average reported in systematic reviews, which found an overall prevalence of 51% (95% CI 36– 66%).(5) However, this finding aligns more closely with data from sub-Saharan Africa, exceeding the prevalence reported in Nigeria (59.7%) (2), but lower than that in Ghana (82%).(12) These results suggest that while the prevalence of academic bullying in our study surpasses the global norm, it is consistent with regional trends.

Bullying predominantly occurred during ward rounds (82.9%), clinical meetings (57.9%), and academic seminars (56.8%), consistent with literature indicating that hierarchical settings in medical environments are common contexts for such behavior.(15,16) Multiple forms of bullying were identified, including unfair criticism, verbal aggression, derogatory remarks, and threats or intimidation. Consultants were the most frequently reported perpetrators, aligning with findings from a systematic review where 53.6% of 15,868 respondents identified senior staff as bullies.(1) These observations underscore the influence of entrenched power dynamics within the medical profession on bullying behaviors.(17)

The high prevalence of bullying in our sample population can be attributed to several factors inherent in the medical profession. Hierarchical power dynamics, overwhelming workloads, and a lack of institutional support have been noted in other studies and are evident in our setting.(15)

Bullying often occurs hierarchically, with senior staff perpetrating negative behaviors toward junior colleagues.(16) The Joint Commission has emphasized that healthcare professionals in positions of power commonly exhibit intimidating and disruptive behaviors, highlighting the systemic nature of the issue.(17)

Toxic work cultures—including bullying and discrimination—are significant sources of distress for junior doctors, necessitating urgent institutional interventions. In Sierra Leone, medical professionals face escalating demands, diminishing resources, and staff shortages, factors known to compound psychological distress.(11) These stressors not only increase the risk of being bullied but also exacerbate the situations under which bullying occurs and intensify its negative impact. The absence of structured systems to counteract this culture may explain the high prevalence observed. Further research is needed to elucidate the role of these stressors specifically related to perpetrators of bullying in the medical profession.

### Determinants of bullying in the medical profession

Our study found no significant differences in the incidence of bullying across demographic factors such as gender, age, marital status, designation, or duration of practice. While previous studies suggest a higher incidence of bullying against females (1,18) — and considering the patriarchal context of Sierra Leone—our data did not reflect significant gender differences. This may be due to sample size limitations, reporting biases, or specific workplace dynamics, and aligns with findings from similar studies in the subregion.(12,19) These results underscore the need for further research and qualitative exploration to uncover underlying factors contributing to bullying.

Similarly, our findings deviate from other studies reporting higher odds of bullying among younger and less experienced individuals, attributed to lower status, perceived vulnerability, and power dynamics.(20) Studies have shown that individuals who are separated, divorced, or widowed have higher odds of reporting bullying than married individuals.(21) However, our study found no statistically significant correlation between marital status and reports of bullying.

The lack of statistically significant findings may be due to sample homogeneity and size; a more extensive and diverse sample could provide greater insight into demographic determinants of bullying, highlighting the need for further studies. Given the homogeneity of our sample, exploration of factors such as race-related bullying, which has been shown to lead to profound psychological distress, was not applicable.(18)

### Impact of academic bullying in the medical profession

Academic bullying has profound impacts on the medical profession. The hierarchical nature of medical training can lead to burnout and dissatisfaction among medical students and residents, deterring them from pursuing further specialization or academic careers.(7)This underscores the broader influence of workplace dynamics on healthcare professionals’ career trajectories and well-being. In Sierra Leone, already facing a shortage of specialized medical staff, the negative effects of academic bullying may exacerbate this issue.(11) Research has demonstrated that victims of bullying may become perpetrators themselves, perpetuating a cycle particularly evident in hierarchical structures where each level may bully the one below. (22)

Studies have highlighted the psychological impact of workplace bullying on junior doctors, including its associations with common mental disorders and suicidal ideation. The detrimental effects extend beyond direct victims to colleagues who may be vicariously impacted. Organizational factors, such as climate, culture, leadership, and support, play significant roles in predicting exposure to bullying, emphasizing the need for holistic approaches to address workplace victimization.

Research has also explored the relationship between workplace bullying and employee turnover intentions, as well as negative implications for productivity and teamwork.(23) The psychological and emotional distress caused by bullying affects both personal and professional lives of junior doctors,(24) a critical concern for nations like Sierra Leone grappling with medical professional shortages. While coping mechanisms such as seeking peer support and focusing on personal growth are employed,(25) systemic changes are imperative to address the root causes of bullying in academic settings. Recognizing workplace bullying as a systemic problem necessitates comprehensive solutions to foster a more supportive and respectful work environment.

### Practical implications

To effectively address academic bullying within USLTHC and the broader Sierra Leone healthcare system, a comprehensive, evidence-based approach is necessary. Establishing culturally sensitive anti-bullying policies is imperative to create a safer and more respectful academic environment. Implementing comprehensive training programs for medical staff— focused on recognizing and preventing bullying, promoting respectful communication, and fostering supportive work environments—is essential. Moreover, advocating for authentic leadership that empowers junior doctors, promotes transparent communication, and addresses hierarchical imbalances can substantially contribute to the mitigation of bullying behaviors in healthcare settings.(26)

Confidential reporting channels, such as anonymous hotlines or independent online platforms, are vital for safeguarding individuals and promoting whistleblowing. Enhancing leadership development within the medical hierarchy is also crucial. Effective leadership models in healthcare enhance learning, teaching, and patient care. By fostering ethical leadership principles, healthcare organizations can cultivate a culture of respect, integrity, and accountability.(27)

Ethical leadership profoundly influences healthcare outcomes, including job satisfaction, safety compliance, and reduction of workplace deviance. The positive impact of ethical leadership on job satisfaction enhances service quality, patient satisfaction, and productivity.(28) Ethical leadership improves safety compliance by building trust among healthcare professionals.(29) Fostering a culture of trust and ethical behavior is therefore crucial for promoting positive outcomes in healthcare organizations.

Addressing incivility and unethical behaviors in healthcare settings is essential. Organizations can leverage Ethics Committees and Clinical Ethics Consultation Services to manage incivility and promote ethical practices.(30) Integrating ethical considerations into organizational practices fosters a supportive and respectful work environment, aligning with the need to cultivate ethical leadership skills among healthcare professionals.(31)

Implementing anti-bullying interventions and creating supportive environments through mentorship, coaching, and feedback mechanisms can mitigate the negative impacts of bullying on junior doctors.(6,32) Fostering a culture of respect and support within medical institutions is essential to promoting the well-being and professional development of all healthcare professionals, including junior doctors.(7,33)

### Strengths and limitations

This study represents the first investigation into academic bullying among junior doctors in Sierra Leone. Strengths include the straightforward administration of the survey, facilitated by a well-educated study population and readily accessible participant list.

However, several limitations must be acknowledged. The reliance on self-reported experiences introduces the potential for response bias, including underreporting due to fear of administrative scrutiny. Additionally, there is a lack of a validated instrument for evaluating academic bullying in an African context. The questionnaire was developed based on prior studies and an extensive literature review. Despite these constraints, the findings suggest disturbingly high levels of perceived bullying and mistreatment during training. Results should be interpreted cautiously, and a higher response rate would have been preferable.

## Conclusions

This study revealed a high prevalence of academic bullying among junior doctors at USLTHC, with unfair criticism, verbal aggression, derogatory remarks, and threats or intimidation being the most common forms identified. Consultants and other senior doctors were frequently identified as perpetrators. Bullying most commonly occurred during ward rounds and clinical meetings. Despite the high prevalence, the analysis did not find any socio-demographic factors significantly associated with the likelihood of experiencing bullying.

Academic bullying in medicine profoundly undermines junior doctors’ mental health and professional development, compromising both individual well-being and the quality of patient care. Confronting this pervasive issue within USLTHC and the broader Sierra Leone healthcare system demands a comprehensive, evidence-based strategy. Key steps include conducting context-specific research to inform culturally sensitive anti-bullying policies, implementing extensive training programs, establishing confidential reporting mechanisms, and promoting ethical leadership.

## Declarations

### Availability of data and materials

Data is available upon reasonable request to the corresponding author, Dr. Mohamed B. Jalloh. All authors recognise the value of sharing individual level data. We aim to ensure that data generated from all our research are collected, curated, managed and shared in a way that maximises their benefit.

### Authors’ contributions

All authors, FJ, ATB, AK, MJJ, KA, MMJF, FMF, OTA, AS, AL, and MBJ were involved in the conceptualisation and planning of the study. FJ, AK, MJJ, KA, MMJF and MBJ were involved with conducting the study, with data collection. FJ, AL and MBJ were involved with the analysis and interpretation of data. FJ and MBJ prepared the first draft of the manuscript. All authors, FJ, ATB, AK, MJJ, KA, MMJF, FMF, OTA, AS, AL, and MBJ contributed to subsequent revisions to the manuscript. All authors, FJ, ATB, AK, MJJ, KA, MMJF, FMF, OTA, AS, AL, and MBJ, read and approved the final manuscript.

### Authors’ information (optional)

Not Applicable

